# Clinical Course and Risk Factors for Recurrence of Positive SARS-CoV-2 RNA: A Retrospective Cohort Study from Wuhan, China

**DOI:** 10.1101/2020.05.08.20095018

**Authors:** Jie Chen, Xiaoping Xu, Jing Hu, Qiangda Chen, Fengfeng Xu, Hui Liang, Nanmei Liu, Hengmei Zhu, Jinlong Lan, Lan Zhou, Jiajun Xing, Ning Pu, Zhigang Cai

**Affiliations:** Department of Cardiothoracic Surgery, Naval Medical Center of PLA, Shanghai, 200052, People’s Republic of China; Department of Infectious Disease, Guanggu Branch of Hubei Province Maternity and Childcare Hospital, Hubei, 430073, People’s Republic of China; Department of General Surgery, Zhongshan Hospital, Fudan University, Shanghai, 200032, People’s Republic of China; Department of Special Treatment, Eastern Hepatobiliary Surgery Hospital, Naval Military Medical University, Shanghai, 200438, People’s Republic of China

**Author notes:** These authors contributed equally to this work and shared the co-first authorship (Jie Chen, Xiaoping Xu, Jing Hu, Qiangda Chen). **Corresponding authors:** Zhigang Cai, Department of Cardiothoracic Surgery, Naval Medical Center of PLA, 338 West Huaihai Road, Changning District, Shanghai, 200052, P.R. China; Ning Pu, Department of General Surgery, Zhongshan Hospital, Fudan University, 180 Fenglin Road, Xuhui District, Shanghai, 200032, P.R. China.

**Keywords:** Coronavirus Disease 2019, SARS-CoV-2, recurrence, clinical course, risk factor

## Abstract

**Background:** Coronavirus Disease 2019 (COVID-19), caused by the severe acute respiratory syndrome coronavirus 2 (SARS-CoV-2), has developed into a full-blown global pandemic. It has been reported that patients with COVID-19 meeting the criteria for hospital discharge (including two consecutive negative RT-PCR results) have experienced recurrent PCR positivity. However, the clinical course and risk factors for these patients have not been well described.

**Methods:** In this retrospective cohort study, consecutive patients with COVID-19 confirmed by RT-PCR from the Guanggu Branch of Hubei Province Maternity and Childcare Hospital from February 24, 2020 to March 31, 2020 were enrolled. All patients received follow-up to April 15, 2020 from discharge. The epidemiological, radiographic, laboratory, treatment, and outcome data were extracted from medical records. Univariate and multivariable logistic regression methods were used to elucidate risk factors for patients with recurrence of positive SARS-CoV-2 RNA.

**Results:** 1087 COVID-19 patients were included in this study. Of these, 20 (1.8%) died and 1067 (98.2%) were discharged from the hospital. Among the discharged cases, there were 81 (7.6%) patients found to develop a repeat positive SARS-Cov-2 RNA result. Older age was obviously associated with death. For patients with recurrent RT-PCR positivity, the median duration from illness onset to onset of complete RNA negative was 33.0 days (range, 6.0-82.0 days; IQR, 20.0-41.0 days), while that from illness onset to recurrence was 50.0 days (range, 21.0-95.0 days; IQR, 36.5-59.5 days). Multivariate regression analysis identified recurrence of positive SARS-Cov-2 RNA was associated with elevated IL-6 levels (P=0.004, OR=3.050; 95% CI, 1.432-6.499), increased lymphocyte count (P=0.038, OR=2.321; 95% CI, 1.048-5.138) and CT imaging features of lung consolidation (P=0.038, OR=1.641; 95% CI, 1.028-2.620) during hospitalization.

**Conclusion:** Elevated lymphocyte counts and IL-6 levels in blood, and consolidation features on CT imaging are useful risk factors for clinicians to identify patients at risk of developing recurrent positivity of SARS-CoV-2 RNA. This is speculated to be caused by a balance in immune regulation when fighting virus toxicity. For patients with a high risk of recurrent positivity, a prolonged observation and additional preventative measures should be implemented for at least 50 days after illness onset to prevent future outbreaks.

**Key Points:** *Question:* How is the clinical course of patients with recurrence of positive SARS-CoV-2 RNA and what clinical characteristics are associated with that?

*Findings:* In this cohort involving 1067 COVID-19 patients discharged from the hospital, 81 (7.6%) patients found to develop a repeat positive SARS-Cov-2 RNA result. For patients with recurrent RT-PCR positivity, the median duration from illness onset to onset of complete RNA negative was 3.30 days (range, 6.0-82.0 days; IQR, 20.0-41.0 days), while that from illness onset to recurrence was 50.0 days (range, 21.0-95.0 days; IQR, 36.5-59.5 days). Risk factors associated with recurrence of positive SARS-Cov-2 RNA included elevated IL-6 levels, increased lymphocyte count and CT imaging features of lung consolidation during hospitalization.

*Meaning:* The recurrence of positive SARS-Cov-2 RNA is speculated to be caused by a balance in immune regulation when fighting virus toxicity. For patients with a high risk of recurrent positivity, a prolonged observation and additional preventative measures should be implemented for at least 50 days after illness onset to prevent future outbreaks.

## Introduction

Coronavirus Disease 2019 (COVID-19), caused by the severe acute respiratory syndrome coronavirus 2 (SARS-CoV-2), was first reported in Wuhan, China and has been spreading globally.^1,2^ As of April 18, 2020, there have been 2,121,675 confirmed cases of COVID-19 and 142,299 related deaths from a total of 213 different countries according to the World Health Organization (WHO).^3^ Undoubtedly, COVID-19 has caused a global pandemic. Thus far, many studies have reported and summarized the epidemiological and clinical features of patients infected with SARS-CoV-2.^4-6^ Furthermore, the pathogenicity and mechanism of SARS-CoV-2 are being wrestled at full stretch and the mystery of SARS-CoV-2 is gradually being unraveled. However, the exact origin species that carried SARS-CoV-2 remains a controversial issue, which is a potential threat to a new outbreak.^7,8^ As we learn more about the origins and course of this disease, we must appropriately looking into the mechanisms of its eradication. Until now, very little know about how the human body regulates and clears a SARS-CoV-2 infection which, in turn, makes it difficult to assess a complete recovery with no risk of infectivity to others. This is essential to halt the COVID-19 spread, “flatten the curve” and prevent additional outbreaks.

In the early stages of the COVID-19 outbreak located in Wuhan, China, the severe shortage and limitations in the detection and accuracy of the RT-PCR test was severely problematic in identifying infected patients. Thankfully, this has since improved drastically with the support of national medical teams from every other provinces comprising of nationwide medical experts and nurses.^9^ This experience gave meaningful insight into the false negative rates of RT-PCR tests and possibility of recurrence of positive SARS-CoV-2 RNA. To counteract the potential probability of false negative rate of RT-PCR tests, patients have been routinely undergone two or more multipoint throat-swabs over 24 hours apart before discharge.^10^ Lan L *et al*.^11^ reported that four medical professionals with COVID-19 who met criteria for hospital discharge (including two consecutive negative RT-PCR results) experienced repeat RT-PCR positivity, implying potential asymptomatic carrier states. Although it has not been demonstrated that patients with recurrent SARS-CoV-2 RNA positive remain infectious after discharge, this is an inevitable matter that must be addressed. Furthermore, our understanding of clinical and radiological characteristics of the patients with COVID-19 who experience recurrence of positive SARS-CoV-2 RNA is very limited.

Herein, we report on 1087 patients with confirmed COVID 19 and further explore a population with repeat positivity of SARS-CoV-2 RNA on RT-PCR during post-hospital isolation and after at least two negative RT-PCR tests from one hospital in Wuhan, China. We aim to present outcomes on this large sample and provide further insight into a unique and understudied population by exploring their clinical course and risk factors.

## Methods

### Study design and participants

This is a retrospective analysis of 1087 consecutive COVID-19 pneumonia patients at the Guanggu Branch of Hubei Province Maternity and Childcare Hospital in Wuhan, China diagnosed by SARS-CoV-2 RNA detection in accordance with the World Health Organization interim guidance. Since the COVID-19 pneumonia outbreak first developed in Wuhan, the military from Naval Medical Center of PLA have completely assumed governance over the Guanggu Branch of Hubei Province Maternity and Childcare Hospital to treat the local COVID-19 pneumonia patients. According to hospital record, all enrolled patients were discharged or died between February 24, 2020 and March 31, 2020. This study was approved by the Research Ethics Committee of Guanggu Branch of Hubei Province Maternity and Childcare Hospital and was granted with a waiver of informed consent from study participants.

### Data collection

An experienced team of front-line medical personnel reviewed and collected the epidemiological, radiographic, laboratory, treatment, and outcome data from medical records to establish a database for COVID-19 pneumonia patients. All patients received follow-up to April 15, 2020 from discharge. The confidential information of patients was protected by assigning a new specific record number. All collected data were checked by two authors (JC and QC) and finally adjudicated by a third researcher (NP) for any differences in interpretation.

### Procedures

In concordance with standard procedure, throat-swab specimens were obtained and tested using real-time RT-PCR methods to identify SARS-CoV-2 infection.^12^ The Academy of Military Medical Sciences and the hospital laboratory were responsible for SARS-CoV-2 detection in respiratory specimens. During the hospital stay and after clinical remission of symptoms, SARS-CoV-2 PCR re-examination by throat-swab specimens was performed at 24-hour intervals. During hospitalization, regular laboratory blood examinations were performed comprising of complete blood counts (including white blood cells, neutrophils, lymphocytes, monocytes and platelets), serum biochemical tests (including liver function tests, renal function indicators and electrolytes), coagulation indices, high-sensitivity C-reactive protein (CRP), erythrocyte sedimentation rate (ESR), procalcitonin, myocardial enzymes, D-dimer and interleukin-6 (IL-6). At an appropriate time determined by the attending physician, computed tomography (CT) scans were routinely performed for inpatients.

The criteria for discharge were as follows: 1) no fever for at least three days; 2) remission of clinical respiratory symptoms; 3) substantial improvement of pulmonary inflammation on chest CT scan; 4) two negative SARS-CoV-2 RNA tests at least 24 hours apart; 5) good general condition.

The illness severity of COVID-19 was defined according to the Chinese management guideline for COVID-19 (version 6.0).^10^ Fever was defined as axillary temperature of at least 37.3°C. Comorbid conditions included hypertension, diabetes and other internal visceral dysfunction during hospitalization (including hypoproteinaemia, coagulopathy, hyperuricemia, anemia, respiratory failure, liver injury, renal injury and cardiac injury).

Hypoproteinaemia was defined as blood albumin of less than 25 g/L, coagulopathy was defined as a 3-second increase in prothrombin time or a 5-second increase of activated partial thromboplastin time, and hyperuricemia was defined as blood trioxypurine greater than 420 umol/L (male) or 360 umol/L (female). Anemia was determined according to WHO guidelines.^13^ Acute respiratory distress syndrome (ARDS) was diagnosed according to the Berlin Definition.^14^ Acute liver failure was diagnosed according to EASL Clinical Practical Guidelines^15^, acute kidney injury was diagnosed according to the KDIGO clinical practice guidelines^16^, and acute cardiac injury was diagnosed as previously reported.^6^

### Statistical analysis

Descriptive analyses of continuous and categorical variables were presented as a median with interquartile range (IQR) and counts with column percentages, respectively. The differences between recurrence and non-recurrence were compared using the Pearson Chis-quared test, Fisher’s exact test or Mann-Whitney U test as appropriate. To explore the risk indicators associated with recurrence of positive SARS-Cov-2 RNA, univariate and multivariate logistic regression models were implemented. Variables with P value <0.2 were selected for multivariable analysis on the basis of previous findings and clinical constraints. Missing data was not imputed and presented as is in Table 4 and 5, and analyses regarding different indicators were based on non-missing data. A two-sided P value less than 0.05 was considered statistically significant. All statistical analyses were performed using the SPSS v21.0 software (IBM Corporation, Armonk, NY, USA), and figures were plotted by GraphPad Prism 8.0 software (GraphPad Software, La Jolla, CA, USA).

## Results

### Demographics and Characteristics of Patients with COVID-19 Pneumonia

A total of 1087 consecutive COVID-19 pneumonia patients with positive SARS-CoV-2 RNA were enrolled in this study. In this cohort, the median patient age was 60.0 years ranging from 9.0 to 100.0 years (IQR, 49.0-69.0 years) and 635 (58.4%) were women. The proportion of general cases was 83.1%, and that of severe and critical cases were 13.2% and 3.7%, respectively. Other general features, inpatient laboratory examinations and imaging findings were shown in **Table 1**. Most of patients (874, 80.4%) had bilateral pulmonary infiltration on chest CT, while 730 patients (67.2%) had a ground-glass appearance, and 525 patients (48.3%) had features of consolidation. 887 out of 1007 (88.1%) patients revealed positive serum IgG, while 797 out of 1057 (75.4%) patients had positive serum IgM for COVID 19.

**Table 1.**
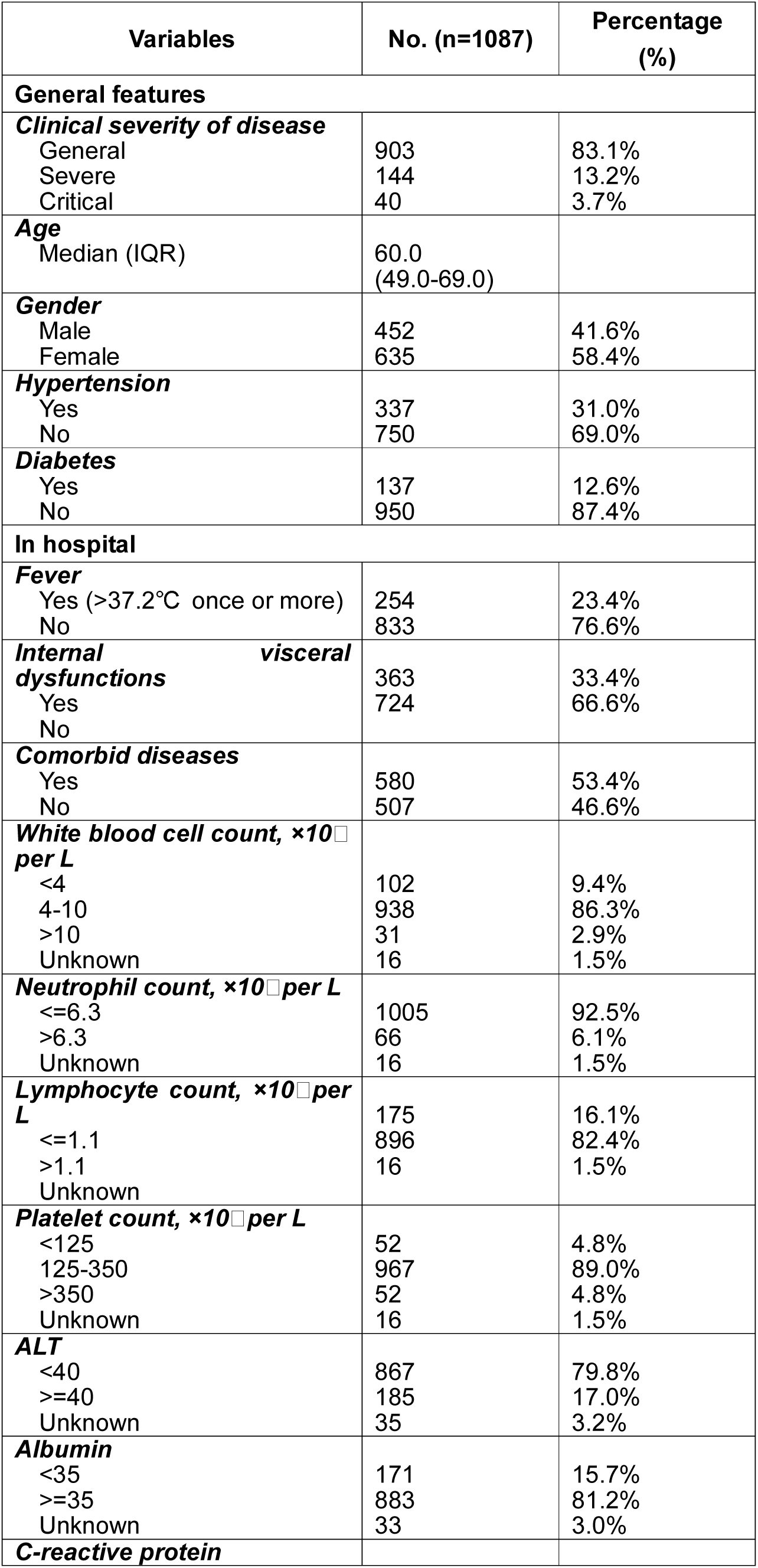
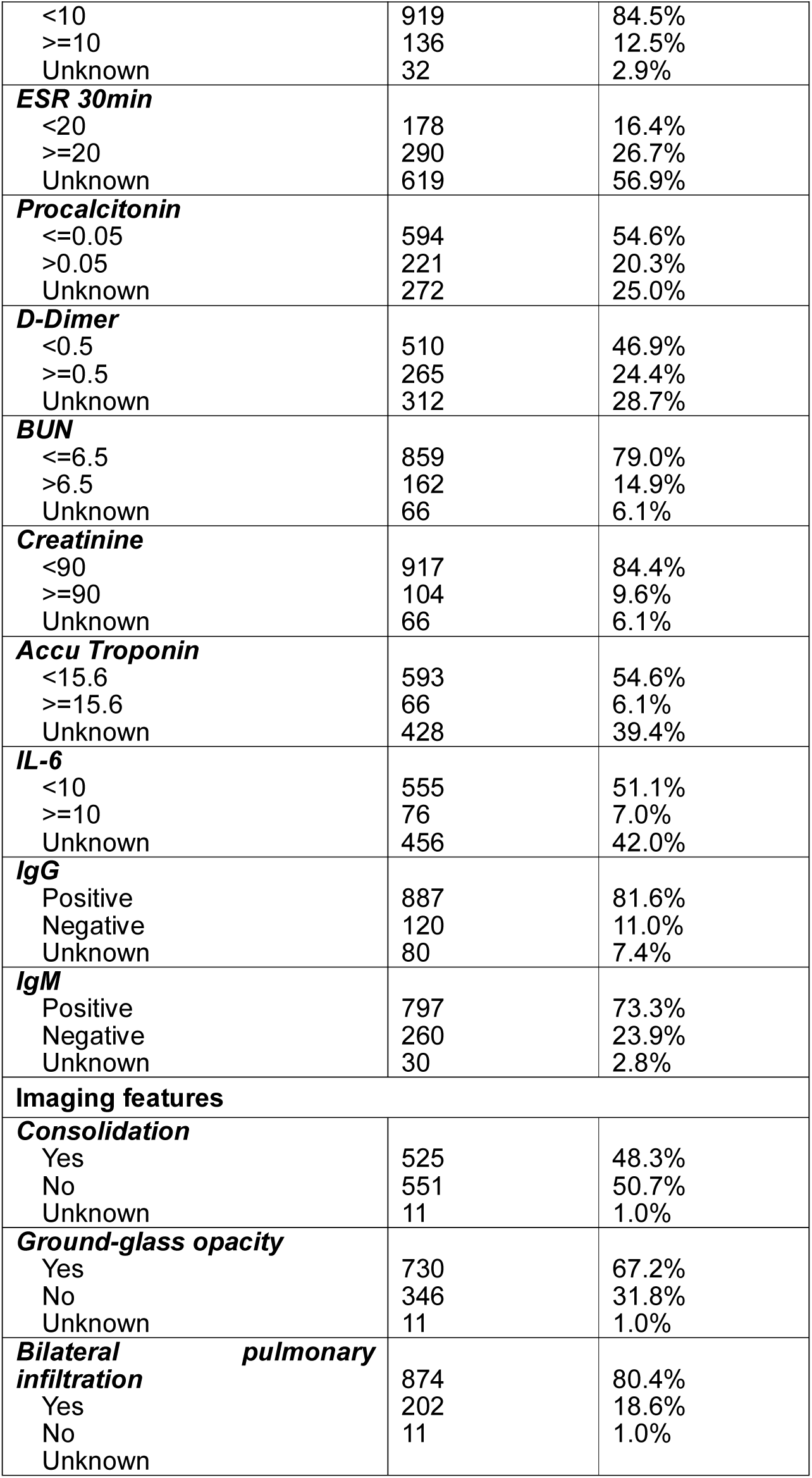
Clinical characteristics of all patients with COVID-19 confirmed by RT-PCR.

| **Variables** | **No. (n=1087)** | **Percentage (%)** |
| --- | --- | --- |
| **General features** |  |  |
| ***Clinical severity of disease*** |  |  |
| General | 903 | 83.1% |
| Severe | 144 | 13.2% |
| Critical | 40 | 3.7% |
| ***Age*** |  |  |
| Median (IQR) | 60.0<br>(49.0-69.0) |  |
| ***Gender*** |  |  |
| Male | 452 | 41.6% |
| Female | 635 | 58.4% |
| ***Hypertension*** |  |  |
| Yes | 337 | 31.0% |
| No | 750 | 69.0% |
| ***Diabetes*** |  |  |
| Yes | 137 | 12.6% |
| No | 950 | 87.4% |
| **In hospital** |  |  |
| ***Fever*** |  |  |
| Yes (>37.2°C once or more) | 254 | 23.4% |
| No | 833 | 76.6% |
| ***Internal visceral dysfunctions*** |  |  |
| Yes | 363 | 33.4% |
| No | 724 | 66.6% |
| ***Comorbid diseases*** |  |  |
| Yes | 580 | 53.4% |
| No | 507 | 46.6% |
| ***White blood cell count, $\times 10^9$ per L*** |  |  |
| <4 | 102 | 9.4% |
| 4-10 | 938 | 86.3% |
| >10 | 31 | 2.9% |
| Unknown | 16 | 1.5% |
| ***Neutrophil count, $\times 10^9$ per L*** |  |  |
| $\leq 6.3$ | 1005 | 92.5% |
| >6.3 | 66 | 6.1% |
| Unknown | 16 | 1.5% |
| ***Lymphocyte count, $\times 10^9$ per L*** |  |  |
| $\leq 1.1$ | 175 | 16.1% |
| >1.1 | 896 | 82.4% |
| Unknown | 16 | 1.5% |
| ***Platelet count, $\times 10^9$ per L*** |  |  |
| <125 | 52 | 4.8% |
| 125-350 | 967 | 89.0% |
| >350 | 52 | 4.8% |
| Unknown | 16 | 1.5% |
| ***ALT*** |  |  |
| <40 | 867 | 79.8% |
| $\geq 40$ | 185 | 17.0% |
| Unknown | 35 | 3.2% |
| ***Albumin*** |  |  |
| <35 | 171 | 15.7% |
| $\geq 35$ | 883 | 81.2% |
| Unknown | 33 | 3.0% |
| ***C-reactive protein*** |  |  |
| <10 | 919 | 84.5% |
| >=10 | 136 | 12.5% |
| Unknown | 32 | 2.9% |
| **ESR 30min** |  |  |
| <20 | 178 | 16.4% |
| >=20 | 290 | 26.7% |
| Unknown | 619 | 56.9% |
| **Procalcitonin** |  |  |
| <=0.05 | 594 | 54.6% |
| >0.05 | 221 | 20.3% |
| Unknown | 272 | 25.0% |
| **D-Dimer** |  |  |
| <0.5 | 510 | 46.9% |
| >=0.5 | 265 | 24.4% |
| Unknown | 312 | 28.7% |
| **BUN** |  |  |
| <=6.5 | 859 | 79.0% |
| >6.5 | 162 | 14.9% |
| Unknown | 66 | 6.1% |
| **Creatinine** |  |  |
| <90 | 917 | 84.4% |
| >=90 | 104 | 9.6% |
| Unknown | 66 | 6.1% |
| **Accu Troponin** |  |  |
| <15.6 | 593 | 54.6% |
| >=15.6 | 66 | 6.1% |
| Unknown | 428 | 39.4% |
| **IL-6** |  |  |
| <10 | 555 | 51.1% |
| >=10 | 76 | 7.0% |
| Unknown | 456 | 42.0% |
| **IgG** |  |  |
| Positive | 887 | 81.6% |
| Negative | 120 | 11.0% |
| Unknown | 80 | 7.4% |
| **IgM** |  |  |
| Positive | 797 | 73.3% |
| Negative | 260 | 23.9% |
| Unknown | 30 | 2.8% |
| **Imaging features** |  |  |
| **Consolidation** |  |  |
| Yes | 525 | 48.3% |
| No | 551 | 50.7% |
| Unknown | 11 | 1.0% |
| **Ground-glass opacity** |  |  |
| Yes | 730 | 67.2% |
| No | 346 | 31.8% |
| Unknown | 11 | 1.0% |
| **Bilateral pulmonary infiltration** |  |  |
| Yes | 874 | 80.4% |
| No | 202 | 18.6% |
| Unknown | 11 | 1.0% |

The median length of hospitalization was 12.0 days (range, 1.0-38.0 days; IQR, 8.0-17.0 days), and 20 patients died during hospitalization, while 1067 were discharged. The total mortality was 1.8% and discharge rate was 98.2%. Within these fatalities, 5 patients were graded as severe cases where the mortality was 3.5%, and 15 were labeled critical cases where the mortality rose to 37.5%. The total mortality of severe and critical cases was 10.6%. The median age of death was 83.0 years ranging from 65.0 to 92.0 years (IQR, 79.3-87.8 years). All of these patients died from multiple organ failure (MOSF), most commonly from lungs, heart, liver and kidneys.

### Characteristics of Patients with Recurrence of Positive SARS-Cov-2 RNA

Among the discharged cases, there were 81 (7.6%) patients found to develop a repeat positive SARS-Cov-2 RNA result during their post-discharge isolation and after two negative RT-PCR tests to warrant initial discharge. In these recurrent cases, the median age was 62.0 years (range, 16-90 years; IQR, 50.5-68.0 years), and 51 (63.0%) patients were female. Twenty (24.7%) patients had comorbid disease of hypertension and 9 (11.1%) patients had diabetes. Divided by clinical severity, general, severe and critical cases accounted for 84.0% (68 cases), 14.8 (12 cases) and 1.2% (1 case), respectively. Most of these patients had initial symptoms before initial COVID-19 diagnosis. However, 15 (18.5%) patients were asymptomatic when first diagnosed by positive SARS-Cov-2 RNA. Before hospitalization, 70 patients were confirmed to harbor pulmonary infection via CT scan, and 65 (65.3%) patients have received therapeutic anti-viral agents.

Laboratory and CT imaging results from the inpatient hospital-stay are summarized in **Table 2**. Of these patients, 7 (8.6%) patients had lymphocytopenia, and only 4 (4.9%) patients had neutrophilia. High sensitivity CRP was elevated in 8 (9.9%) patients, increased ESR was found in 27 (33.3%) patients and increased procalcitonin in 14 (17.3%) patients. Additionally, the increased inflammatory factor of IL-6 was found in 11 (13.6%) patients. Regarding organ dysfunction, 10 (12.3%) patients developed liver injury with elevated ALT, 4 (4.9%) patients demonstrated myocardial damage with elevated accu troponin, and 11 (13.6%) patients incurred kidney injury with elevated serum BUN and creatinine. Characteristics of inpatient CT images revealed consolidation, ground-glass opacity and bilateral pulmonary infiltration in 49 (60.5%), 56 (69.1%) and 70 (86.4%) patients, respectively. 72 out of 77 (93.5%) patients manifested positive serum IgG, while 68 out of 79 (86.1%) patients had positive serum IgM for COVID-19.

**Table 2.**
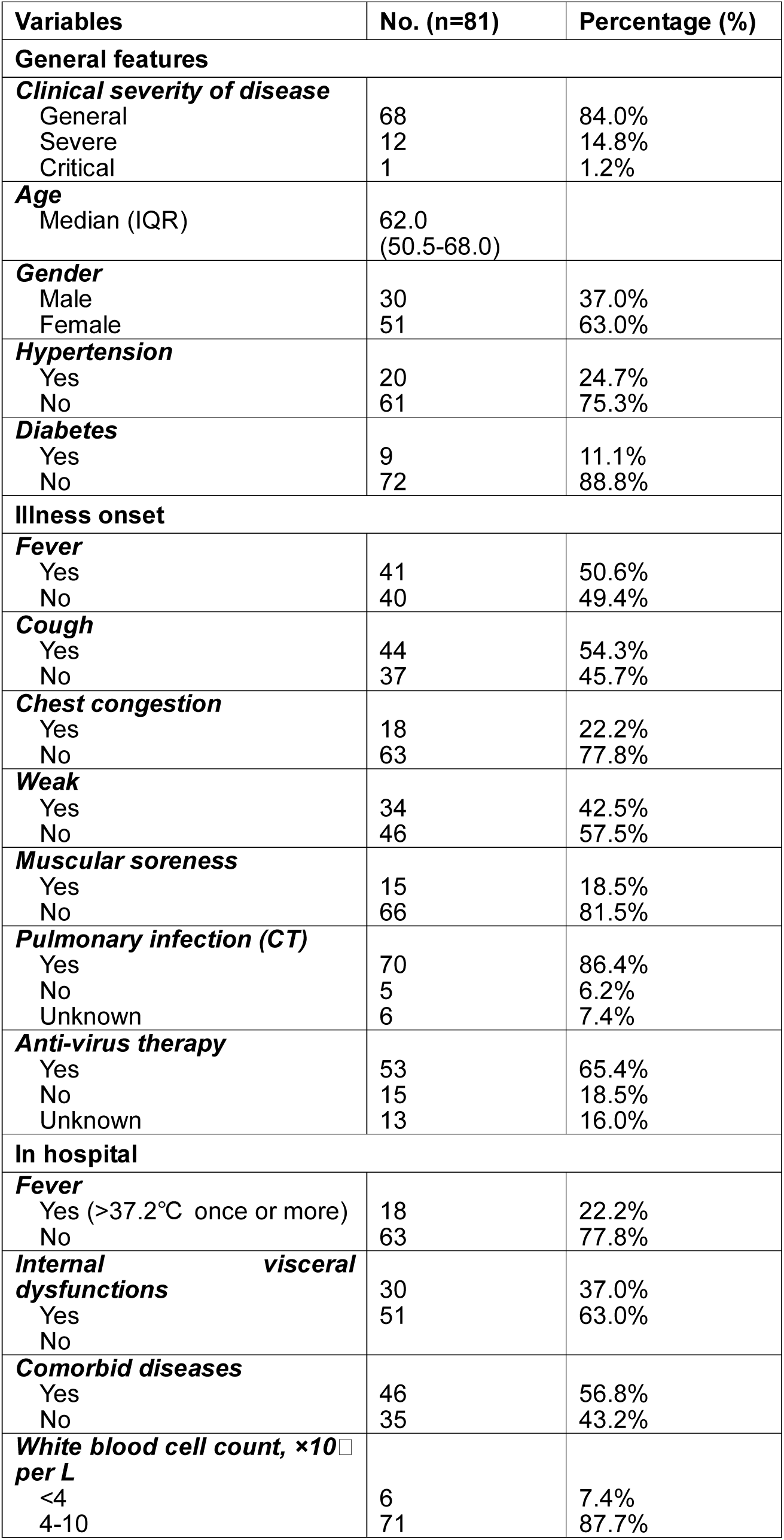
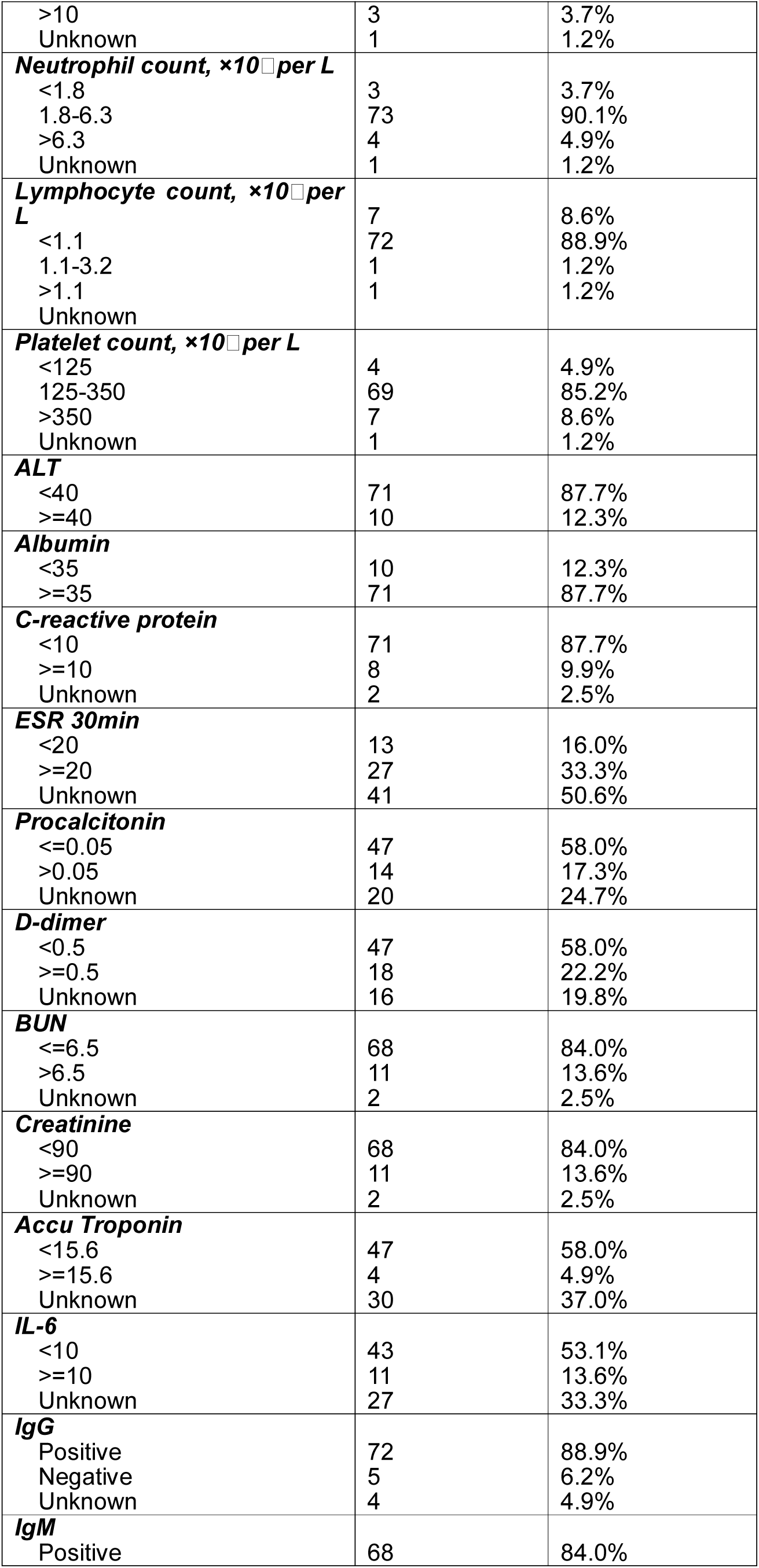
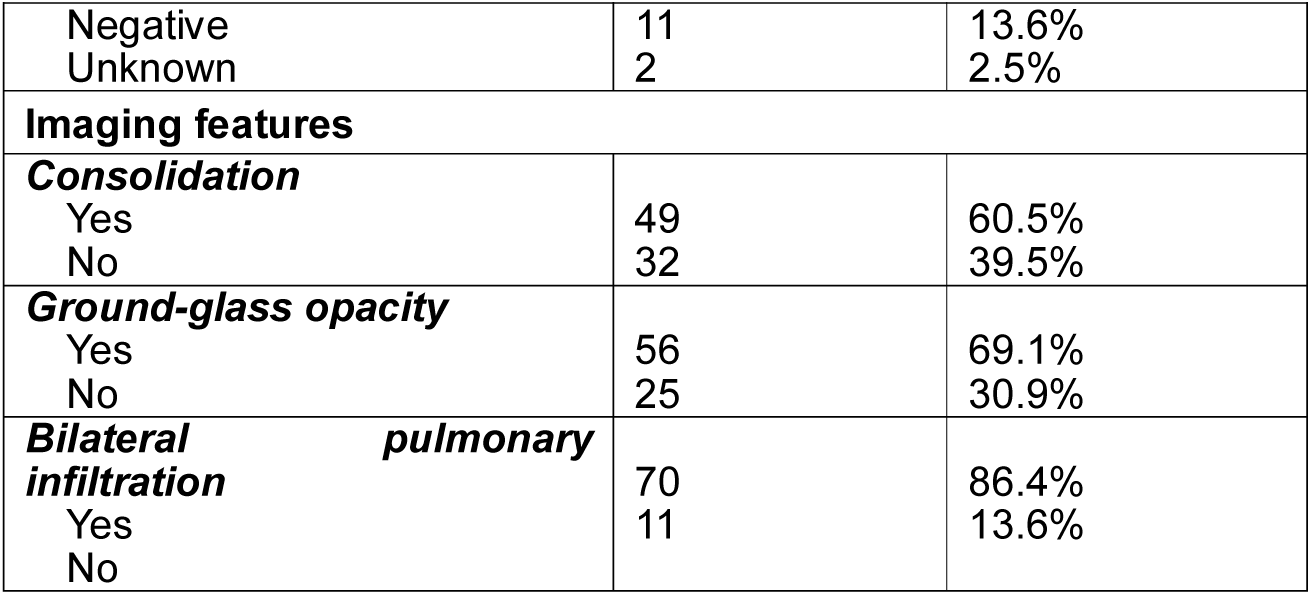
Clinical characteristics of patients with recurrence of positive SARS-Cov-2 RNA.

### Clinical Course of Patients with Recurrence of Positive SARS-Cov-2 RNA

As depicted in **Figure 1**, the median length of hospitalization for patients with recurrence of positive SARS-Cov-2 RNA was 12.0 days (range, 4.0-27.0 days; IQR, 7.0-17.0 days). From **Figure 2A**, the median length from discharge to recurrence was 9.0 days (range, 3.0-18.0 days; IQR, 7.0-10.0 days), the median duration for these patients from illness onset to RT-PCR confirmation if COVID-19 was 11.0 days (range, 0-57.0 days; IQR, 1.5-21.0 days), and from illness onset to onset of complete RNA negative was 33.0 days (range, 6.0-82.0 days; IQR, 20.0-41.0 days), while that from illness onset to recurrence was 50.0 days (range, 21.0-95.0 days; IQR, 36.5-59.5 days). Then from **Figure 2B**, the median duration from initial diagnostic RT-PCR to recurrence was 36.0 days (range, 16.0-64.0 days; IQR, 26.5-45.0 days). In addition, the median duration between initial diagnostic RT-PCR and onset of complete RNA negative was 17.0 days (range, 1.0-45.0 days; IQR, 8.0-29.0 days), while between onset of complete RNA negative and recurrence it was 12.0 days (range, 4.0-27.0 days; IQR, 7.0-17.0 days).

**Figure 1.**
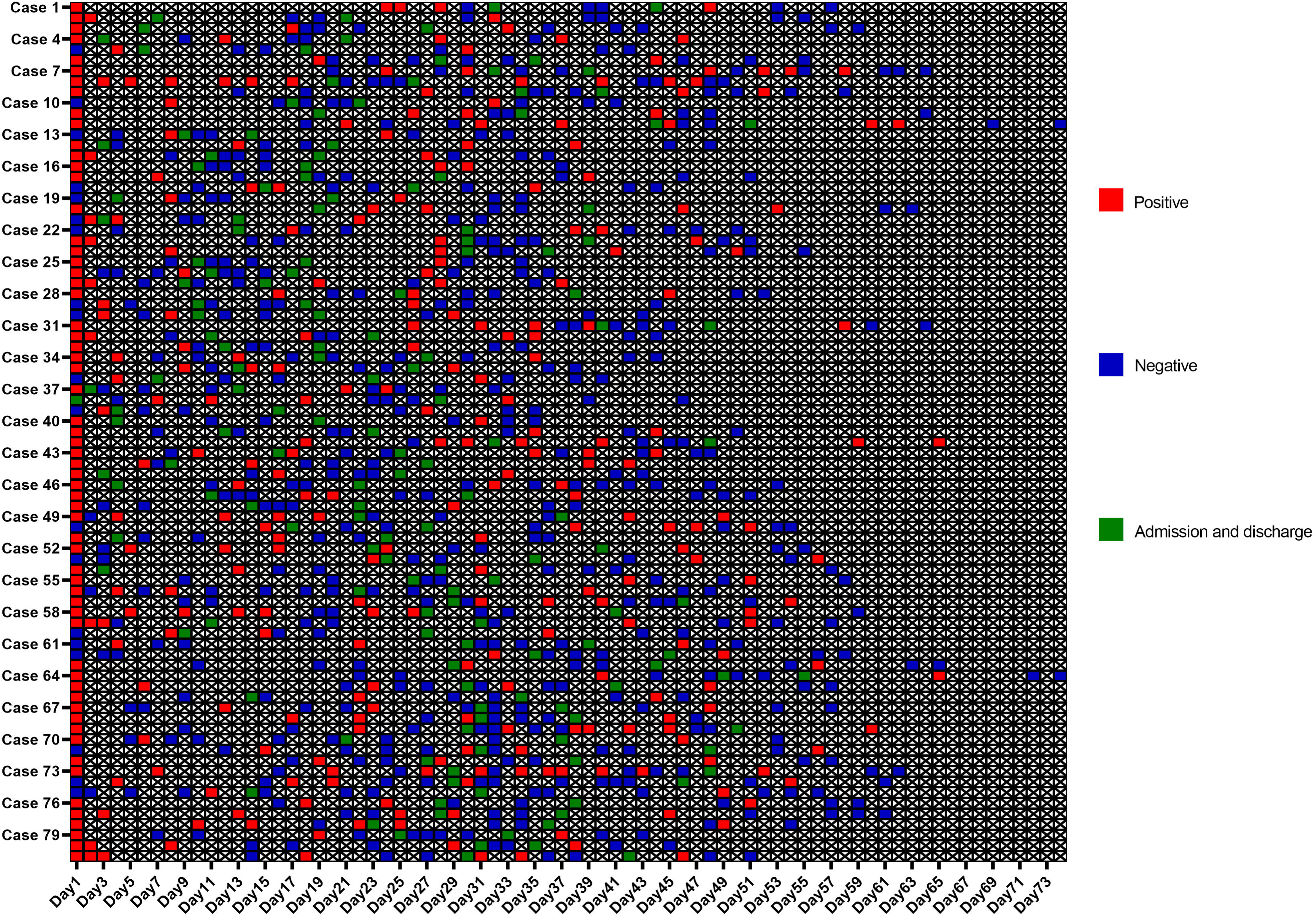
Individual duration of viral shedding and recurrence from illness onset to repeat positivity in patients with recurrence of positive SARS-CoV-2 RNA after discharge. Figure shows the timing and results of RT-PCR examinations for SARS-CoV-2 RNA in details. SARS-CoV-2=severe acute respiratory syndrome coronavirus 2. RT-PCR=reverse transcription-polymerase chain reaction.

**Figure 2.**
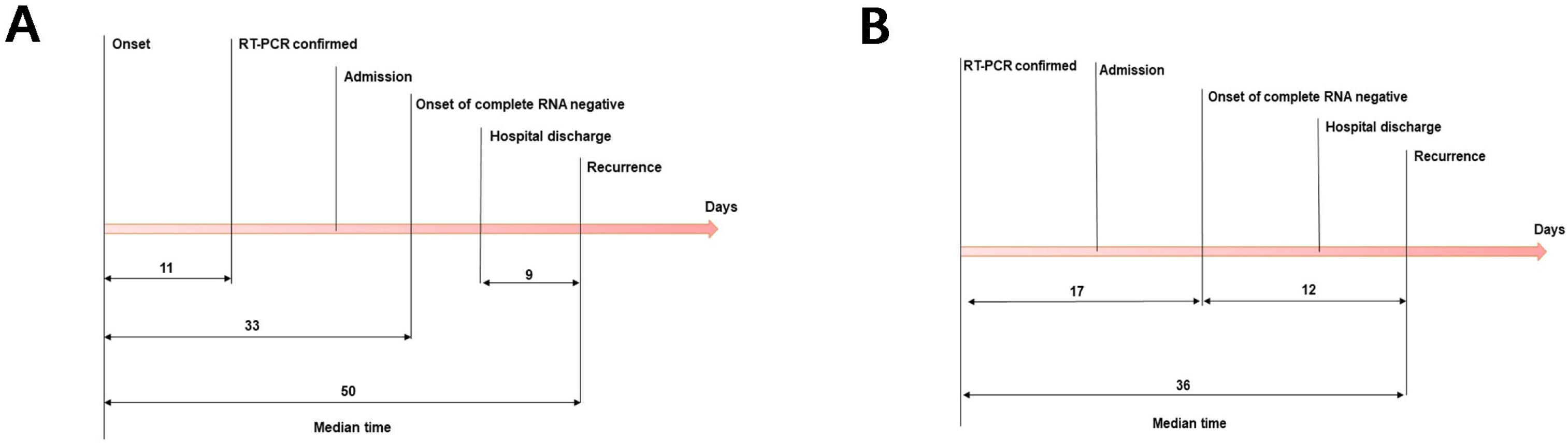
The median duration of different stages in patients with recurrence of positive SARS-CoV-2 RNA after discharge. Figure 2A shows the median duration from illness onset to initial RT-PCR confirmed, onset of complete RNA negative and recurrent RT-PCR positivity after discharge, and from discharge to recurrence. Figure 2B shows the median duration from initial RT-PCR confirmed to onset of complete RNA negative and recurrent RT-PCR positivity after discharge, and from onset of complete RNA negative to recurrence. SARS-CoV-2=severe acute respiratory syndrome coronavirus 2. RT-PCR=reverse transcription-polymerase chain reaction.

Amongst these 81 patients, oxygen support was administrated in 37 (45.7%), however, no invasive mechanical ventilation (IMV) or IMV with extracorporeal membrane oxygenation (ECMO) was used. The optimal antiviral therapy was administrated in 69 (85.2%) patients; these included abidor hydrochloride (40 patients, 49.4%), interferon alfa (17 patients, 21.0%), entecavir/tenofovir (7 patients, 8.6%) and oseltamivir (5 patients, 6.2%). More than half of patients (51, 63.0%) were treated with Chinese patent drugs, such as Lianhuaqingwen capsule. Vitamin C was given to 41 (50.6%) patients, and immunomodulators, such as thymopentin and immunoglobulin, were administrated in 8 (9.9%) patients.

### Associated Risk Factors with Recurrence of Positive SARS-CoV-2 RNA

As shown in **Table 3**, significant positive correlations were found between recurrence of positive SARS-CoV-2 RNA and serum IL-6 level (P=0.010) and CT imaging depicting consolidation (P=0.031). In the univariate analysis, elevated lymphocyte count (P=0.194, OR=1.644; 95% CI, 0.776-3.484), elevated serum IL-6 level (P=0.013, OR=2.504; 95% CI, 1.218-5.150), consolidation on CT imaging (P=0.033, OR=1.655; 95% CI, 1.042-2.629) and bilateral pulmonary infiltration (P=0.196, OR=1.540; 95% CI, 0.800-2.966) were considered as potential risk factors for recurrence of SARS-Cov-2 RNA positivity (**Table 4**). Multivariate analysis concluded that elevated lymphocyte count (P=0.038, OR=2.321; 95% CI, 1.048-5.138), serum IL-6 level (P=0.004, OR=3.050; 95% CI, 1.432-6.499) and consolidation features on CT imaging (P=0.038, OR=1.641; 95% CI, 1.028-2.620) remained as independent risk predictors for recurrence (**Table 5**).

**Table 3.** Correlations between clinical characteristics and recurrence of positive SARS-Cov-2 RNA in discharged patients.

| Variables | No.<br>(n=1067) | No recurrence<br>n=986 | Recurrence<br>n=81 | P value |
| --- | --- | --- | --- | --- |
| **General features** |  |  |  |  |
| **Clinical severity of disease** |  |  |  | 0.684 |
| General | 903 | 835 | 68 |  |
| Severe | 139 | 127 | 12 |  |
| Critical | 25 | 24 | 1 |  |
| **Age** |  |  |  | 0.700 |
| Median (IQR) | 60.0<br>(49.0-68.0) | 60.0<br>(49.0-68.0) | 62.0<br>(50.5-68.0) |  |
| **Gender** |  |  |  | 0.424 |
| Male | 440 | 410 | 30 |  |
| Female | 627 | 576 | 51 |  |
| **Hypertension** |  |  |  | 0.200 |
| Yes | 331 | 311 | 20 |  |
| No | 736 | 675 | 61 |  |
| **Diabetes** |  |  |  | 0.664 |
| Yes | 135 | 126 | 9 |  |
| No | 932 | 860 | 72 |  |
| **In hospital** |  |  |  |  |
| **Fever** |  |  |  | 0.853 |
| Yes (>37.2°C once or more) | 246 | 228 | 18 |  |
| No | 821 | 758 | 63 |  |
| **Internal visceral dysfunctions** |  |  |  | 0.327 |
| Yes | 343 | 313 | 30 |  |
| No | 724 | 673 | 51 |  |
| **Comorbid diseases** |  |  |  | 0.419 |
| Yes | 560 | 514 | 46 |  |
| No | 507 | 472 | 35 |  |
| **White blood cell count, $\times 10^9$ per L** |  |  |  | 0.579 |
| <4 | 100 | 94 | 6 |  |
| 4-10 | 927 | 856 | 71 |  |
| >10 | 24 | 21 | 3 |  |
| Unknown | 16 | 15 | 1 |  |
| **Neutrophil count, $\times 10^9$ per L** |  |  |  | 1.000 |
| $\leq 6.3$ | 994 | 918 | 76 | |
| >6.3 | 57 | 53 | 4 |  |
| Unknown | 16 | 15 | 1 |  |
| **Lymphocyte count, $\times 10^9$ per L** |  |  |  | 0.190 |
| $\leq 1.1$ | 158 | 150 | 8 | |
| >1.1 | 894 | 821 | 72 |  |
| Unknown | 15 | 15 | 1 |  |
| **Platelet count, $\times 10^9$ per L** |  |  |  | 0.297 |
| <125 | 44 | 40 | 4 |  |
| 125-350 | 956 | 886 | 69 |  |
| >350 | 52 | 45 | 7 |  |
| Unknown | 15 | 15 | 1 |  |
| **ALT** |  |  |  | 0.202 |
| <40 | 852 | 781 | 71 |  |
| $\geq 40$ | 181 | 171 | 10 | |
| Unknown | 34 | 34 | 0 |  |
| **Albumin** |  |  |  | 0.502 |
| <35 | 154 | 144 | 10 |  |
| $\geq 35$ | 880 | 809 | 71 | |
| Unknown | 33 | 33 | 0 |  |
| **C-reactive protein** |  |  |  | 0.653 |
| <10 | 914 | 843 | 71 |  |
| >=10 | 121 | 113 | 8 |  |
| Unknown | 32 | 30 | 2 |  |
| **ESR 30min** |  |  |  | 0.420 |
| <20 | 176 | 163 | 13 |  |
| >=20 | 282 | 255 | 27 |  |
| Unknown | 609 | 568 | 41 |  |
| **Procalcitonin** |  |  |  | 0.571 |
| <=0.05 | 589 | 542 | 47 |  |
| >0.05 | 207 | 193 | 14 |  |
| Unknown | 271 | 251 | 20 |  |
| **D-dimer** |  |  |  | 0.339 |
| <0.5 | 507 | 460 | 47 |  |
| >=0.5 | 250 | 232 | 18 |  |
| Unknown | 310 | 294 | 16 |  |
| **BUN** |  |  |  | 0.822 |
| <=6.5 | 853 | 785 | 68 |  |
| >6.5 | 148 | 137 | 11 |  |
| Unknown | 66 | 64 | 2 |  |
| **Creatinine** |  |  |  | 0.150 |
| <90 | 907 | 839 | 68 |  |
| >=90 | 94 | 83 | 11 |  |
| Unknown | 66 | 64 | 2 |  |
| **Accu Troponin** |  |  |  | 1.000 |
| <15.6 | 590 | 543 | 47 |  |
| >=15.6 | 54 | 50 | 4 |  |
| Unknown | 423 | 393 | 30 |  |
| **IL-6** |  |  |  | **0.010** |
| <10 | 552 | 509 | 43 |  |
| >=10 | 63 | 52 | 11 |  |
| Unknown | 452 | 425 | 27 |  |
| **Imaging features** |  |  |  |  |
| **Consolidation** |  |  |  | **0.031** |
| Yes | 520 | 471 | 49 |  |
| No | 541 | 509 | 32 |  |
| Unknown | 6 | 6 | 0 |  |
| **Ground-glass opacity** |  |  |  | 0.798 |
| Yes | 720 | 664 | 56 |  |
| No | 341 | 316 | 25 |  |
| Unknown | 6 | 6 | 0 |  |
| **Bilateral pulmonary infiltration** |  |  |  | 0.193 |
| Yes | 859 | 789 | 70 |  |
| No | 202 | 191 | 11 |  |
| Unknown | 6 | 6 | 0 |  |

**Table 4.** Univariate regression analysis for risk factors of patients with recurrence of positive SARS-Cov-2 RNA.

| Variables | No. | Univariate OR<br>(95% CI) | P value |
| --- | --- | --- | --- |
| Age | 1067 | 1.000(0.985-1.015) | 0.995 |
| Gender | 1067 | 1.210(0.757-1.933) | 0.425 |
| Clinical severity of disease | 1067 | 0.976(0.579-1.645) | 0.927 |
| Hypertension | 1067 | 0.712(0.422-1.200) | 0.202 |
| Diabetes | 1067 | 0.853(0.416-1.749) | 0.665 |
| Fever | 1067 | 0.950(0.551-1.637) | 0.853 |
| Internal visceral dysfunctions | 1067 | 1.265(0.790-2.025) | 0.328 |
| Comorbid diseases | 1067 | 1.207(0.764-1.906) | 0.420 |
| Neutrophil count, $\times 10^9$ per L | 1051 | 0.912(0.321-2.587) | 0.862 |
| Lymphocyte count, $\times 10^9$ per L | 1051 | 1.644(0.776-3.484) | **0.194** |
| Platelet count, $\times 10^9$ per L | 1051 | 1.417(0.676-2.969) | 0.356 |
| ALT | 1033 | 0.643(0.325-1.273) | 0.205 |
| Albumin | 1034 | 1.264(0.637-2.508) | 0.503 |
| C-reactive protein | 1035 | 0.841(0.394-1.792) | 0.653 |
| ESR 30min | 458 | 1.328(0.666-2.647) | 0.421 |
| Procalcitonin | 796 | 0.837(0.450-1.553) | 0.572 |
| D-dimer | 757 | 0.759(0.431-1.337) | 0.340 |
| Accu Troponin | 644 | 0.924(0.320-2.671) | 0.884 |
| IL-6 | 615 | 2.504(1.218-5.150) | **0.013** |
| Consolidation | 1061 | 1.655(1.042-2.629) | **0.033** |
| Ground-glass opacity | 1061 | 1.066(0.653-1.740) | 0.798 |
| Bilateral pulmonary infiltration | 1061 | 1.540(0.800-2.966) | **0.196** |

**Table 5.**
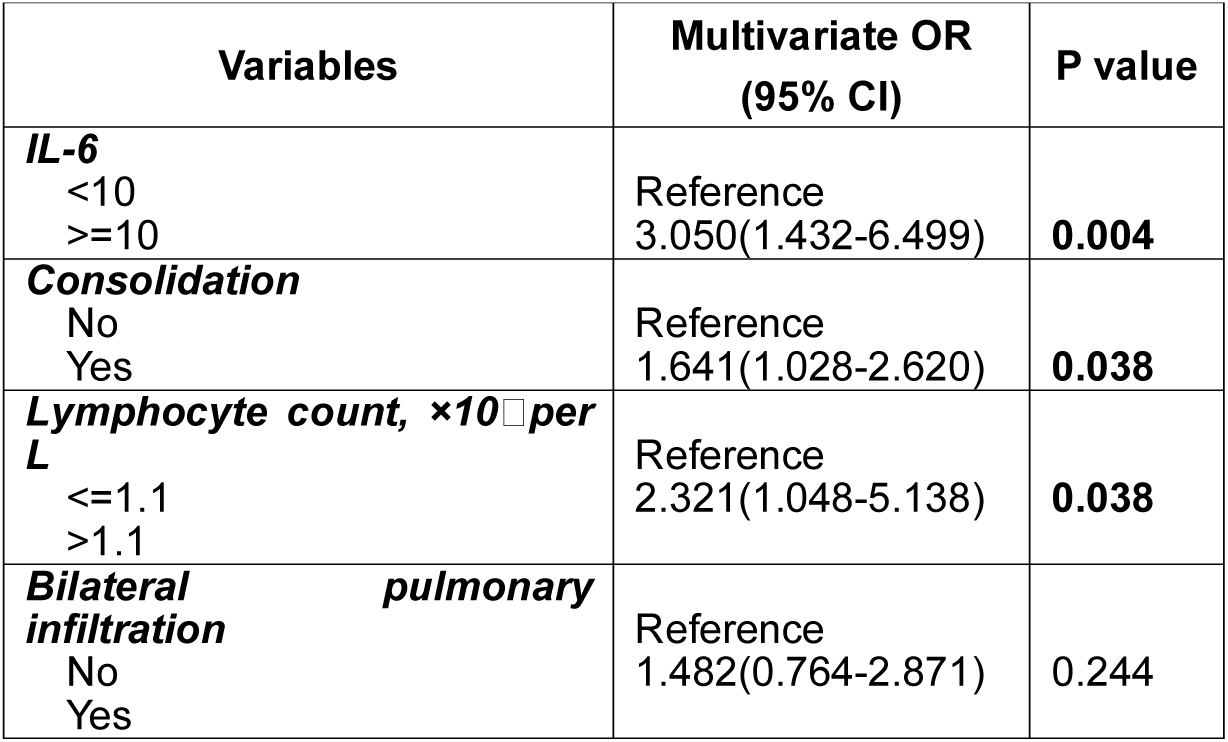
Multivariate regression analysis for risk factors of patients with recurrence of positive SARS-Cov-2 RNA.

| **Variables** | **Multivariate OR**<br>**(95% CI)** | **P value** |
| --- | --- | --- |
| **IL-6**<br><10<br>≥10 | Reference<br>3.050(1.432-6.499) | **0.004** |
| **Consolidation**<br>No<br>Yes | Reference<br>1.641(1.028-2.620) | **0.038** |
| **Lymphocyte count, ×10<sup>9</sup> per L**<br>≤1.1<br>>1.1 | Reference<br>2.321(1.048-5.138) | **0.038** |
| **Bilateral pulmonary infiltration**<br>No<br>Yes | Reference<br>1.482(0.764-2.871) | 0.244 |

## Discussion

In this study, we provide comprehensive data on the demographic and clinical characteristics of 1087 consecutive patients with COVID-19 from Wuhan, China. According to the degree of severity of COVID 19, the enrolled patients were categorized as 903 (83.1%) general cases, 144 (13.2%) severe cases and 40 (3.7%) critical cases on admission. The total mortality of severe and critical cases was 10.6%. The mortality of the all patients with COVID-19 pneumonia was 1.8%, which was similar to that of a previous study^4^, but lower than that reported in another studies.^5,17^ This difference may be partly attributed to the fact that the majority proportion of cases were classified as general severity and that more medical resources were made available in the later stages of this pandemic which matches our window of enrollment into this study. Liang WH *et al*,^18^ reported that the mortality of COVID-19 patients out of the Hubei Province was limited to 0.3%, as strict public health interventions were initiated in order to prevent further outbreak outside Hubei and medical resources were adequately provided for treatment. Additionally, previous studies indicated that older age was an important independent predictor for mortality in COVID-19 patients.^6,12^ Our findings support this claim; the median age of death was 83.0 years, distinctly higher than that of the discharged patients, which further suggests that increased age was associated with an increased risk of mortality.

Of 1067 discharged patients, we identified 81 (7.6%) patients that experienced recurrence of positive SARS-Cov-2 RNA after meeting criteria for discharge. This is similar to findings in previous case reports.^11,19,20^ However, Yuan J *et al.^20^* reported a higher repeat positivity rate of 14.5% after discharge, which may be due to a small cohort of enrolled patients. This group of patients may pose a potential risk for further disease spread as persistent asymptomatic viral carriers. This not so rare phenomenon threatens potential future outbreaks of COVID-19 that seems to have been temporarily controlled in China after a series of unprecedented public health interventions.^21^ Thus, we aimed to explore the clinical course and risk predictors for recovered COVID-19 patients with recurrent PCR positivity to provide insight into this population and help guide clinical practice in order to help halt future outbreaks.

Various studies have reported on infectivity and viral shedding. Zhou F *et al.^6^* showed that the median duration of viral shedding was 20.0 days in survivors and the longest observed duration was 37 days. Moreover, Zhou B *et al.^22^* reported that the median duration of viral shedding was days from illness onset in severe COVID-19 patients. Xu K *et al.^23^* reported that almost 3 in every 4 patients had viral RNA clearance within 21 days of illness onset, and male gender, older age, hypertension, delayed hospital admission after illness onset, severe illness upon admission, invasive mechanical ventilation and corticosteroid treatment were risk factors for extended viral RNA clearance. In our study, the median duration of viral shedding for patients with recurrence of positive SARS-CoV-2 RNA was 33.0 days from illness onset to onset of complete RNA negative. However, the median duration from illness onset to recurrent SARS-CoV-2 RNA positive was 50.0 days. Thus, our findings underscore the importance of a prolonged treatment or isolation for patients at increased risk of recurrence of SARS-CoV-2 RNA positivity.

Amongst the 81 discharged patients with recurrence of positive SARS-CoV-2 RNA, we found that age and comorbid diseases, which were previously described to be risk factors for mortality^12^, were not significant risk factors when compared with patients with no recurrence. However, three independent predictors were identified for patients with recurrence of positive SARS-Cov-2 RNA after treatment and hospital discharge. Elevated serum IL-6 level, lymphocyte count greater than 1.1*10^8^ /L and consolidation on CT imaging during hospitalization were associated with higher likelihoods of recurrent SARS-Cov-2 RNA positivity after discharged. Partly similar to previous study, it showed that lymphocyte concentrations before discharge were significantly positively correlated with the time interval for virus reappearing, which confirmed the role of lymphocytes in the potential recurrence of SARS-CoV-2 RNA positivity.^20^ No significant differences were found in other factors in our cohort, including clinical severity of disease, CRP, D-dimer level, etc. IL-6, one of the main pro-inflammatory factors of the immune system, plays an important role in host defense against infections. However, due to the ability of SARS-CoV-2 to infect the lower respiratory tract and rapidly replicated leads to excessive IL-6 release inducing an acute severe systemic inflammatory response known as cytokine release syndrome (CRS).^24^ Previously, increased serum IL-6 level was reported to be observed in severe and critical patients with COVID-19 and to be associated with poor outcomes^25,26^, which is a similar finding during severe acute respiratory syndrome (SARS) outbreak.^27^ Concurrently, lymphopenia was also commonly noted in patients with COVID-19, especially in severe and critical cases^5,26,28^, suggesting dysregulated immune responses in this sub-cohort. However in our study, only 175 (16.1%) of 1087 cases showed a decrease in lymphocyte count, which again may be due to the fact that the cohort is largely comprised of general cases. Interestingly, we discovered that the discharged patients with recurrence of positive SARS-CoV-2 RNA may potentially have an elevated serum IL-6 level and lymphocyte count than those with no recurrence, implying that the immune responses against SARS-CoV-2 might still be attempting to clear the infection. Perhaps the immune system could suppress but not eradicate SARS-CoV-2 completely, which may have led to the false-negative results due to lower viral loads. Theoretically, at some point further down the line, the virus started replicating again to cause recurrent positive RT-PCR test results in the already discharged patients with COVID-19.

The characteristics of chest CT imaging features of COVID-19 pneumonia are very useful for preliminary judgment and have contributed to a lower rate of missed diagnoses.^29^ Patients with features of consolidation on CT imaging were reported to associated with more critical cases.^30^ Progression of consolidation might represent further infiltration of the lung parenchyma and lung interstitium, indicating that the virus has invaded the respiratory epithelium which is characterized by diffuse alveolar damage and necrotizing bronchitis. This leads to alveoli being completely filled by inflammatory exudate.^31,32^ Thus, during the recovery process of COVID-19 patients with lung consolidation, a potentially undetectable amount of SARS-CoV-2 may persist in the respiratory epithelium. This may result in the recurrence of positive SARS-CoV-2 RNA after discharge. Interestingly, we found that most patients with recurrence of positive SARS-CoV-2 RNA had fluctuating positive and negative results in the course of the disease, typically in cases 7, 8, 41 amongst others (**Figure 1**). This, in itself, may partially reflect another potential sign for recurrent positivity after discharge. Also, such fluctuations in one case partly ruled out the randomly error probability in RT-PCR detection. Thus, individuals may have already had immunity to eradicate the virus, so a period of duration was needed for complete recovery yet. However, if the individual immunity cannot deal with the recurrence, further treatment may be still needed.

### Limitations

This study has a few notable limitations. First, this study was conducted at a single-center hospital. As such, there may be an element of selection bias where identifying factors may influence the clinical outcomes. A larger cohort study of patients with COVID-19 pneumonia from different institutions nationwide or worldwide would help to further define the clinical characteristics and risk factors of recurrence. Second, only multipoint of throat-swab specimens were performed for patients in this study. Thus the chance of false negative results is still possible. Multisite sampling could be collected for RT-PCR detection, such as the fecal SARS-CoV-2 RNA test, particularly in patients with gastrointestinal symptoms.^33^

## Conclusions

Elevated lymphocyte counts and IL-6 level in blood, and consolidation on chest CT were associated with a greater risk of developing recurrent positivity of SARS-CoV-2 RNA, possibly due to a balance between immune regulation and virus toxicity. For patients with a higher risk of recurrent positivity, a prolonged treatment or isolation period for observation should be carried out for at least 50 days after illness onset in order to identify patients who may pose a risk for future outbreaks.

## Data Availability

Data will be made available when requested

## Author Contributions

Drs Cai and Pu had full access to all of the data in the study and take responsibility for the integrity of the data and the accuracy of the data analysis. Joint first authors are Drs J. Chen, X. Xu, Hu, and Q. Chen.

Concept and design: J. Chen, Pu, Cai.

Acquisition, analysis, or interpretation of data: All authors.

Drafting of the manuscript: J. Chen, Q. Chen, Pu, Cai.

Critical revision of the manuscript for important intellectual content: J. Chen, X. Xu, Hu, Q. Chen, Pu, Cai.

Statistical analysis: J. Chen, Q. Chen, Liu, Pu, Cai.

Obtained funding: J. Chen.

Administrative, technical, or material support: J. Chen, F. Xu, Pu, Cai.

Supervision: Pu, Cai.

## Additional Contributions

We acknowledge all health-care workers involved in the diagnosis and treatment of patients in Wuhan; and we thank Joseph R Habib (Johns Hopkins University School of Medicine, Baltimore, Maryland, USA) for review and revise of the manuscript.

